# Robust data-driven auditory profiling towards precision audiology

**DOI:** 10.1101/2020.04.09.20036442

**Authors:** Raul Sanchez-Lopez, Michal Fereczkowski, Tobias Neher, Sébastien Santurette, Torsten Dau

## Abstract

The sources and consequences of a sensorineural hearing loss are diverse. While several approaches have aimed at disentangling the physiological and perceptual consequences of different etiologies, hearing deficit characterization and rehabilitation have been dominated by the results from pure-tone audiometry. Here, we present a novel approach based on data-driven profiling of perceptual auditory deficits that attempts to represent auditory phenomena that are usually hidden by, or entangled with, audibility loss. We hypothesize that the hearing deficits of a given listener, both at hearing threshold and at supra-threshold sound levels, result from two independent types of “auditory distortions”. In this two-dimensional space, four distinct “auditory profiles” can be identified. To test this hypothesis we gathered a dataset consisting of a heterogeneous group of listeners that were evaluated using measures of speech intelligibility, loudness perception, binaural processing abilities and spectro-temporal resolution. The subsequent analysis revealed that distortion type-I was associated with elevated hearing thresholds at high frequencies and reduced temporal masking release and was significantly correlated with elevated speech reception thresholds in noise. Distortion type-II was associated with low-frequency hearing loss and abnormally steep loudness functions. The auditory profiles represent four robust subpopulations of hearing-impaired listeners that exhibit different degrees of perceptual distortions. The four auditory profiles may provide a valuable basis for improved hearing rehabilitation, e.g. through profile-based hearing-aid fitting

## Introduction

Currently, “profiling” has gained broad attention as a tool for typifying groups of observations (e.g. users, recordings or patients) that follow similar patterns. Data-driven profiling can uncover complex structures that are “hidden” in the data. It has been used as a diagnostic tool in various fields (Shah, Kendall, Khozin, Goosen, Hu, Laramie, Ringel and Schork 2019) such as functional imaging (Krohne, Wang, Hinrich, Moerup, Chan and Madsen 2019), genetics (Li, Rao, Wang and Gong 2004), psychology (Gerlach, Farb, Revelle and Nunes Amaral 2018) or logopedics (Sharma, Purdy and Humburg 2019). The idea of using computational data analysis that applies principles of the knowledge discovery from databases (KDD; Frawley, Piatetsky-Shapiro and Matheus 1992) has recently gained attention in the field of audiology in connection with hearing-aid features (Lansbergen and Dreschler 2020; Mellor, Stone and Keane 2018). As in stratified medicine (Trusheim, Berndt and Douglas 2007), which pursues the identification of subgroups of patients (phenotypes) for the purpose of implementing more targeted treatments, it is of interest to identify subgroups of hearing-impaired (HI) listeners who might benefit from targeted hearing-aid fittings. As such, data-driven auditory profiling could help identify groups of listeners that are characterized by specific hearing disabilities and support precision audiology.

Hearing devices are the usual treatment for a hearing loss (Cunningham and Tucci 2017). Hearing-aid fitting mainly consists of the adjustment of amplification parameters to compensate for audibility loss and impaired loudness perception. Advanced hearing-aid signal processing features, such as adaptive compression speed, beamforming and noise reduction, are typically not individually adjusted in this process, even though they could, in principle, be considered in the compensation of supra-threshold hearing deficits (Kiessling 2001; Neher, Wagener and Fischer 2016). However, the characterization of individual supra-threshold hearing deficits can be complex and requires more testing than standard audiometry. The definition of supra-threshold auditory deficits is commonly based on Plomp’s (Plomp 1978) model, where hearing deficits affecting speech intelligibility are comprised of an “attenuation” and a “distortion” component. Whereas the attenuation component is assumed to affect speech intelligibility only in quiet, the distortion component is assumed to do so also in noise, yielding elevated speech reception thresholds in both cases. Kollmeier and Kiessling (2018) extended Plomp’s approach and suggested a model that includes an attenuation component (affecting pure-tone sensitivity), a distortion component (affecting speech intelligibility in noise), and a neural component (affecting binaural processing abilities). Their model assumes that a sensorineural hearing loss is characterized by several factors: an “audibility loss”, a “compression loss”, a “central loss” and a “binaural loss”. In general, these modelling approaches (Plomp 1978; Kollmeier and Kiessling 2018) are rather conceptual and do not pinpoint specific underlying impairment factors nor specific measures to quantify these types of losses.

There have been some attempts to stratify HI listeners based on the shapes of their audiograms. Several classification schemes have been proposed in earlier studies, some of which were based on data-driven approaches (Bisgaard, Vlaming and Dahlquist 2010; Chang, Yoon, Kim, Baek, Cho, Hong, Kim and Moon 2019; Parthasarathy, Romero Pinto, Lewis, Goedicke and Polley 2020), where computational methods for data analysis were used for identifying the most common audiometric profiles. Based on results from human temporal bone studies, Schuknecht and Gacek (1993) proposed four different types of age-related hearing loss: sensory presbycusis, neural presbycusis, metabolic presbycusis and mechanical presbycusis. Sensory presbycusis was related to alterations in the Organ of Corti and typically associated with basilar membrane compression loss, reduced frequency selectivity and elevated audiometric thresholds. This type of age-related hearing loss was considered to reflect the loss of outer hair cells (OHC; Ahroon, Davis and Hamemik 1993) and/or inner hair cells (IHC; Lobarinas, Salvi and Ding 2013) and was characterized by sloping audiograms. Neural presbycusis was related to a substantial loss of nerve fibers in the spiral ganglion. This type of presbyacusis was characterized by a progressive loss of speech discrimination performance, even though the audiometric thresholds remained unchanged over the same time period. Metabolic presbycusis was related to the atrophy of the stria vascularis that affects the OHC function and the transduction in the sensory cells because of a decreased endo-cochlear potential (EP). This type of impairment was associated with flat audiograms and did not affect speech discrimination (Pauler, Schuknecht and Thornton 1986). Finally, conductive presbycusis corresponded to a gently sloping hearing loss at high frequencies, not reflecting morphological alterations in the sensory cells or stria vascularis but yielding elevated thresholds. This type of presbyacusis might reflect an atypical organization in the organ of Corti that affects its mechanical properties (Motallebzadeh, Soons and Puria 2018; Raufer, Guinan and Nakajima 2019). However, recent results obtained with new techniques developed for histopathological analysis suggested that OHC dysfunction might have been underestimated in age-related hearing loss (Wu et al. 2020).

Animal studies, where selective damage to the sensory cells or a change of the EP was induced, have allowed a consistent definition of the metabolic and sensory types of impairments in terms of audibility loss (Ahroon et al. 1993; Lobarinas et al. 2013; Mills et al. 2006). Dubno et al. (2013) proposed a classification into sensory and metabolic audiometric phenotypes based on an approach that combined findings from animal models, expert medical advice and data-driven techniques. The main goal of their study was to analyze a large database of audiograms of HI individuals, and to identify connections between the findings from the animal studies with induced hearing losses and those based on human data. Whereas Schuknecht and Gacek (1993) characterized the metabolic and sensory types of presbyacusis in terms of physiological impairments observed in humans, Dubno et al. (2013) proposed a phenotypical classification of the audiograms of HI listeners. Dubno *et al*.’s classification was thus solely based on the shape of the pure-tone audiogram. While this may help predict the possible origin of a listener’s audibility loss, supra-threshold auditory processing deficits cannot be inferred from their phenotypes. The perceptual consequences of sensory or metabolic presbyacusis beyond audibility loss have not yet been studied.

We hypothesize that a listener’s hearing deficit can be characterized by two independent types of “auditory distortions”, type-I and type-II, as illustrated in Figure 1. In this two-dimensional space, a normal-hearing (NH) listener would be placed at the origin whereas other listeners, with auditory deficits that differ in the degree of the two types of distortions, would be placed at different positions along the two dimensions. Each type of distortion would then be defined by specific deficits observed in behavioral tasks that covary together and define a given auditory profile. While Profile C represents a high degree of both types of distortion, profiles B and D reflect hearing deficits dominated by one of the two distortions. Profile A, the group with a low degree of distortions, represents only mild hearing deficits.

**Figure 1.**
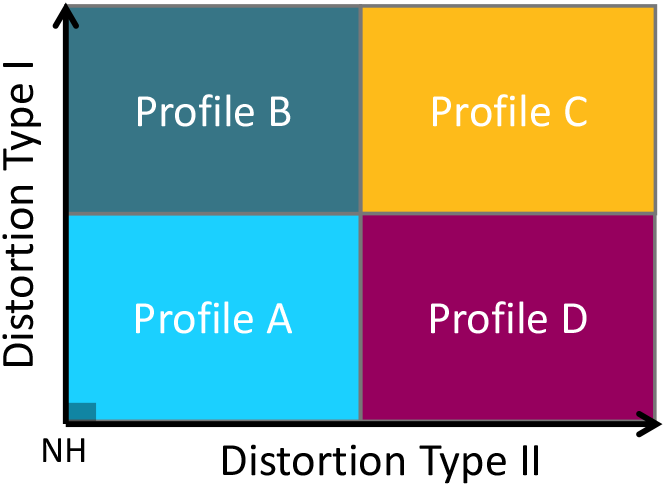
Sketch of the hypothesis. The hearing deficits of a given listener can be described as a combination of two independent perceptual distortions. In a two-dimensional space, there would be four subgroups of listeners (Profiles A-D), which exhibit different degrees of the two distortion types.

Recently, Sanchez-Lopez, Bianchi, Fereczkowski, Santurette and Dau (2018) proposed a data-driven method for auditory profiling that was tested and verified by analyzing two datasets from previous experimental studies (Thorup, Santurette, Jørgensen, Kjærbøl, Dau and Friis 2016)^*^, (Johannesen, Pérez-González, Kalluri, Blanco and Lopez-Poveda 2016) ^†^. The method was tailored to the hypothesis of the four auditory profiles. In their study, it was hypothesized that distortion type-I covaries with a loss of audibility, whereas distortion type-II was assumed to be unrelated to audibility. However, the results of the analysis of two different datasets did not support this hypothesis. In fact, the analysis of the two datasets showed that distortion type-I was connected to high-frequency hearing loss and reduced speech intelligibility. Regarding distortion type-II, the analysis of one of the datasets (Thorup et al. 2016) provided a link to reduced binaural processing abilities, whereas the analysis of the other dataset (Johannesen et al. 2016) was linked to low-frequency hearing loss. These mixed results were attributed to differences between the two datasets in terms of the selection of the listeners and chosen behavioral tests. The authors concluded that a new dataset that included a larger variability of impairment factors across listeners was needed to better characterize the listeners’ auditory distortions and, thus, the auditory profiles. Furthermore, they suggested that the chosen tests should investigate several aspects of auditory processing while at the same time being clinically feasible (i.e. time-efficient, reliable and reasonably accomplishable for patients with diverse abilities).

The current study focused on the scientific basis of the auditory profiling and not on its application in audiological practice. A new dataset was therefore generated with the aim of overcoming the limitations discussed in Sanchez-Lopez et al. (2018). Seventy-five older listeners with different hearing abilities. The behavioural tasks included measures of audibility, loudness perception, binaural processing abilities, speech perception, spectro-temporal modulation sensitivity and spectro-temporal resolution (Sanchez-Lopez et al. 2020). These outcomes include several measures that can be connected to previous approaches, such as the attenuation-distortion model (regarding speech perception measures) and the neural component (regarding binaural processing abilities). Therefore, it was of interest to further investigate the connections between outcome measures and the two distortion types in a data-driven approach. The analysis of the new dataset was performed with a refined version of the data-driven method provided in Sanchez-Lopez et al. (2018). Importantly, the current study did not aim to disentangle the effects of audibility and supra-threshold deficits but to identify four robust listener subpopulations based on the data-driven analysis of the new dataset. The outcomes of the analysis were discussed in relation to previous classification approaches as well as in terms of implications towards profile-based rehabilitation strategies. Moreover, a decision tree consisting of the auditory measures that best classified the listeners into the four profiles was generated.

## Method

The data-driven method was proposed as an alternative to the “expert-driven” method used by Dubno et al. (2013). Although experts in hearing science and audiology can classify listeners based on different criteria, the present approach adopted a hypothesis-driven approach to the data-driven analysis method. Therefore, the development of the data-driven method for auditory profiling was based on two premises: 1) the identification of relevant outcome measures that tap into two independent sources of variation, and 2) the identification of extreme exemplars that can serve as “prototypes” of different subgroups of listeners.

### Description of the dataset

Seventy-five listeners participated in the study. Seventy of the listeners presented various degrees and shapes of symmetrical, sensorineural hearing losses, while five showed normal audiometric thresholds (≤ 25 dB HL in the frequency range between 0.25 and 4 kHz). The participants were aged between 59 and 82 years (median: 71 years). Thirty-eight of them were female. Additionally, one young normal-hearing listener with experience with the tests (participant 0) was included for the analysis as suggested in Sanchez-Lopez et al. (2018). This participant reflected an “optimal performer” and was used as a reference in the profiling method. The listeners were recruited from the clinical databases at Odense University Hospital (OUH), Odense, Denmark and Bispebjerg Hospital (BBH), Copenhagen, Denmark and the Hearing System Section of the Technical University of Denmark (DTU), Kgs Lyngby, Denmark. All listeners completed the “BEAR test battery” (Sanchez-Lopez et al. 2020). This test battery consists of a total of 10 psychoacoustic tests. The tests are divided into six aspects of auditory processing: audibility, speech perception, loudness perception, binaural processing abilities, spectro-temporal modulation sensitivity and spectro-temporal resolution.

The tests were carried out in a double-walled booth (at BBH and DTU) or in a small anechoic chamber (at OUH). The stimuli were presented via headphones (Sennheiser HDA200). The stimuli were presented monaurally, except for those used for testing binaural processing abilities. The stimulus level was adjusted to be audible (i.e. above the audiometric threshold). For example, the test of interaural phase differences was presented at 35 dB sensation level (SL), whereas the tone-in-noise detection task was performed using a noise level of 70 dB HL.^‡^

The dataset (Sanchez-Lopez et al. 2019) consisted of 26 outcome variables corresponding to 75 listeners with different hearing abilities. Table 1 summarizes the outcome variables used in the analysis.

**Table 1.**
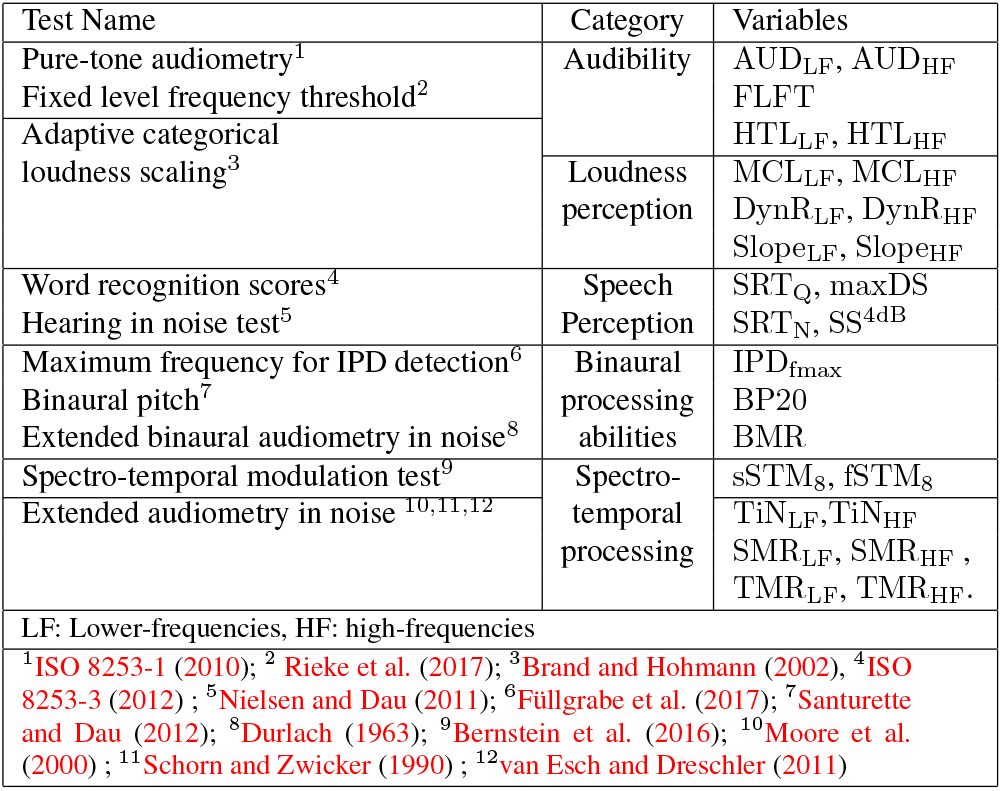
Description of the tests, dimensions and outcome measures contained in the BEAR3 dataset (Sanchez-Lopez et al. 2019). For each test, a reference is included. The tests are divided by categories, and the outcome variables are presented in the right column.

There were six outcomes related to audibility (AUD) and loudness perception (LOUD) represented in a total of 11 variables: 1) pure-tone average at low frequencies (AUD_LF_; f ≤ 1kHz) and at higher frequencies (AUD_HF_; f > 1kHz); 2) fixed-level frequency threshold (FLFT) measured at 80 dB sound pressure level (SPL); 3) hearing threshold levels (HTL) estimated from the loudness function, averaged for low (HTL_LF_) and high (HTL_HF_) frequencies; 4) most comfortable level (MCL) estimated from the loudness function, averaged for low (MCL_LF_) and high (MCL_HF_) frequencies; 5) dynamic range (DynR) estimated as the difference between the uncomfortable level and HTL, estimated from the loudness function for low (DynR_LF_) and high (DynR_HF_) frequencies; and 6) slope of the loudness function at low (Slope_LF_) and high (Slope_HF_) frequencies. For the outcome measures estimated from the loudness function, the low-frequency average corresponded to the center frequencies 0.25, 0.5 and 1 kHz and the high-frequency average corresponded to the center frequencies 2, 4 and 6 kHz. There were four variables related to speech perception. Two of them related to speech-in-quiet (SiQ): 1) speech reception threshold in quiet (SRT_Q_); 2) maximum word recognition score (Max DS); and two of them related to speech-in-noise (SiN): 3) speech reception threshold in noise (SRT_N_); and 4) sentence recognition score at +4 dB SNR (SS_4_^dB^). There were three variables related to binaural processing abilities (BIN): maximum frequency for detecting an interaural phase difference of 180^◦^ (IPD_fmax_); binaural pitch detection performance, estimated as the percent correct of the dichotic presentations (BP_20_); and binaural masking release (BMR). BMR was estimated as the difference between the threshold in the diotic tone-in-noise detection condition (*N*_0_*S*_0_) and the threshold in the dichotic tone-in-noise detection condition where the tone was out of phase between the ears (*N*_0_*S*π). The frequency of the tone presented in the two conditions was 0.5 kHz. The spectrotemporal modulation (STM) and processing (STP) variables included: 1) short-STM test, which assesses spectrotemporal modulation sensitivity at +3 dB modulation depth (sSTM_8_) and 2) the “fast” spectro-temporal modulation detection threshold (fSTM_8_). In both tests, the stimulus was a three-octave wide noise centered at 0.8 kHz which was spectro-temporally modulated Bernstein et al. (2016); 3) the tone-in-noise detection threshold at 500 Hz (TiN_LF_) and at 2 kHz (TiN_HF_). The spectro-temporal processing abilities were assessed by two derived measures: 4) spectral masking release (SMR) estimated as the difference between the tone-in-noise detection threshold (TiN) and the corresponding threshold with the noise shifted towards higher frequencies (center frequency of the noise, *fc,noise* =1.1*ftone*); and 5) temporal masking release (TMR) estimated as the difference between the TiN masked threshold and the corresponding threshold with the tone presented in temporally-modulated noise (modulation frequency, *fm* =4 Hz).

### Pre-processing of the data

For each of the tests, the outcome measures of interest were extracted from the raw results. For example, the speech reception threshold (SRT) in quiet was estimated from the word discrimination scores obtained at different speech levels. When the tests contained frequency-specific measures, the results were grouped into low-frequency (≤ 1 kHz) and high-frequency (*>* 1 kHz) averages. This decision was motivated by previous studies (Bernstein et al. 2016; Sanchez-Lopez et al. 2018; Wu et al. 2019) where a similar division of the averaged audiometric thresholds was undertaken. In the case of monaural measures, the mean values across ears were used. The data were cleaned following the principles of KDD, to remove outliers or unreliable data before the analysis. For example, some of the listeners performed the speech-in-noise test at lower levels than the level recommended for the hearing-in-noise measurements (Nielsen and Dau 2011). Since speech-in-noise perception is of great interest in the present analysis, unreliable measurements of speech reception thresholds in noise (SRT_N_) and sentence recognition scores (SScore_4_*^dB^*) were considered as missing data. In the next step, the data were normalized between the 25th and 75th percentiles, such that the 25th percentile corresponded to a value of - 0.5 and the 75th to a value of 0.5. In total, 26 variables were selected from the outcome measures, as shown in Table 1. The resulting dataset (‘BEAR3’) is publicly available (Zenodo doi:10.5281/zenodo.3459579; Sanchez-Lopez et al. 2019).

### Stages of the data-driven method

As in Sanchez-Lopez et al. (2018), the data-driven analysis used here was based on unsupervised learning and was divided into three main steps illustrated in the top panel of Figure 2:

I. Dimensionality reduction: Based on principal component analysis (PCA), a subset of variables that were highly correlated with the first two principal components, PC1 and PC2, was kept for the following steps (II and III). The subset could consist of 3, 4 or 5 variables per PC. Hence, up to 10 variables could be kept for the next step. The to-be-kept variables were chosen in an iterative process using a leave-one-out cross-validation. In each iteration, one variable was removed according to the variance explained by the remaining variables i.e., the subset of variables that explained the largest amount of variance was kept and the left-out variable was discarded. Additionally, since the use of several intercorrelated variables in PCA can bias the results, highly correlated variables were removed. If two variables resulting from step I were highly correlated (Pearson’s correlation coefficient, r > 0.85), one of them was dropped and this step was repeated.
II. Archetypal analysis: The data were decomposed into two matrices – the ‘test matrix’, which contained the extreme patterns of the data (archetypes) and the ‘subject matrix’, which contained the weights for each archetype. A given subject was then represented as a convex combination of the archetypes (Cutler and Breiman 1994). The specific method used here was similar to the one proposed in Mørup and Hansen (2012). The analysis was limited to four archetypes to improve the interpretability of the results on the scope of the hypothesis.
III. Profile identification: The subject matrix was used to estimate the distance between observations and the four archetypes. Each listener (subject) was then assigned to an auditory profile group based on their weights in the subject matrix. The sum of weights for each listener was always 1. Listeners with a weight above 0.51 for one of the four archetypes were identified as belonging to that auditory profile (Ragozini et al. 2017). Otherwise, they were left “unidentified” (‘U’).

**Figure 2.**
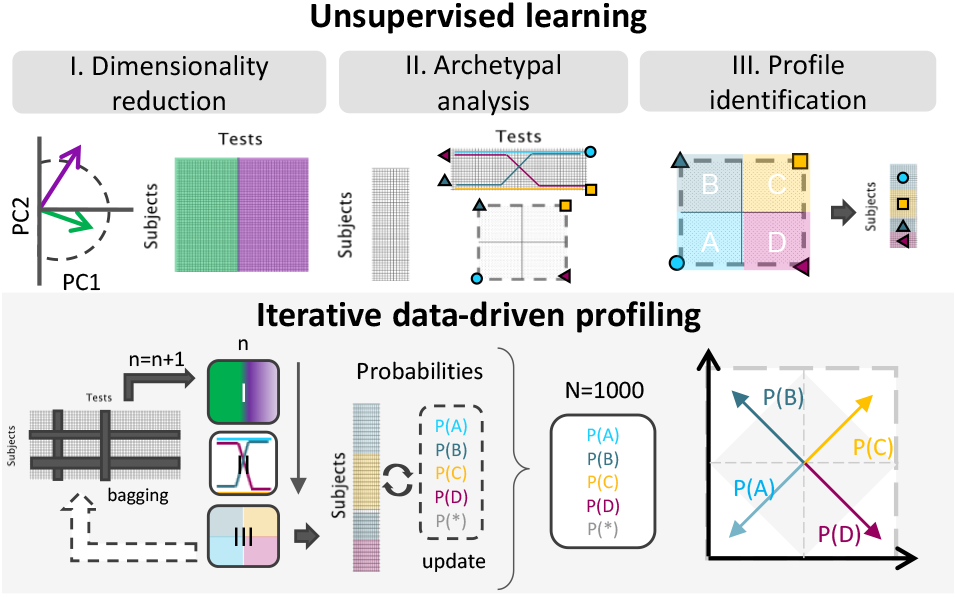
Sketch of the refined data-driven method for auditory profiling. Top panel: The unsupervised learning stages of Sanchez-Lopez et al. (2018): I) dimensionality reduction; II) archetypal analysis; III) profile identification. Bottom panel: In each iteration, a subset of the dataset was processed using dimensionality reduction, archetypal analysis and profile identification. The profile identification stage was two-fold: 1) In each of the iterations, the profiles were identified based on the archetypal analysis. 2) After 1000 iterations, the probability was calculated based on the prevalence of each observation and the number of identifications as each of the profiles. Listeners with higher probabilities of belonging to an auditory profile were placed close to the corners in the square representations and the ones with lower probability (P < 0.5) were located inside the grey square.

### Iterative data-driven profiling

The robust data-driven auditory profiling method aimed to improve the previous method proposed in Sanchez-Lopez et al. (2018) by reducing the influence of the data on the definition of the auditory profiles. In any data-driven analysis, and especially in unsupervised learning, individual data points can clearly influence the results and lead to misinterpretations. Resampling techniques such as bagging are commonly used for supervised learning. Moreover, bagging can improve cluster analysis, making the results less sensitive to the type and number of variables (Dudoit and Fridlyand 2003). The three unsupervised learning steps were repeated 1000 times, as illustrated in the bottom panel of Figure 2. Before each repetition, the full dataset was decimated randomly in terms of subjects and tests in each iteration. The analysis was performed with only 83% of the data (69 out of 75 listeners and 24 out of 26 variables) in each repetition. In the case of missing data, an algorithm based on spring metaphor was used to predict those data points. Furthermore, in step I (dimensionality reduction), the number of selected variables (6, 8 or 10) was also randomly selected in each iteration to further randomize the procedure. Steps II (archetypal analysis) and III (profile identification) yielded a pre-classification of the subjects contained in the subset of the data corresponding to each iteration. The probability of each listener of being identified as a given auditory profile depended on the number of times a given listener was “out-of-bag” in individual repetitions and the profile identification result from step III. In each iteration, the profile probabilities [P(A), P(B), P(C) or P(D)] and the probability of being unidentified [P(U)] were updated. This iterative process was chosen to avoid that few individual listeners bias the derived profiles.

After 1000 repetitions, the listeners were divided into four subgroups based on the computed probabilities. If a given listener showed a probability above 0.5 of belonging to any of the auditory profiles, the listener was assigned to that profile. However, if the highest probability was below 0.5, but P(U) was also below 0.5, the listener was considered “in-between” two profiles. The criterion for the “in-between” listeners to be included in one of the four clusters was that the difference between the two highest probabilities had to be above 0.1 to be considered significant. The remaining of the “in-between” listeners were considered inconclusive and no assigned to any profile. The projection of the probabilities on a two-dimensional space was done by considering four vectors, one for each profile probability, pointing towards each of the corners in a squared representation, as depicted in the right-bottom panel of Figure 2. Graphically, the listeners belonging to an auditory profile were then placed close to the corners.

### Distortion estimation from the square representation

The final output of the refined data-driven method was the probability, P, of being identified as belonging to an auditory profile (A-D). Regarding the square representation or convex hull, which resembled the hypothesis shown in Figure 1, the probabilities of belonging to an auditory profile were depicted as vectors with the origin at the center of the square and oriented towards each of the four corners (Figure 2). Assuming that P(B) and P(C) are proportional to auditory distortion type-I (AD_I_) and that assuming that P(C) and P(D) are proportional to auditory distortion type-II (AD_II_), this yields:

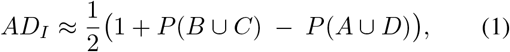

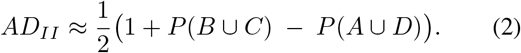

Each listener was placed in the two-dimensional space along with the two estimated distortions. In addition, prototypes reflecting extreme exemplars, equivalent to the archetypes yielded by the archetypal analysis, were estimated by averaging the results of the five listeners with the highest probabilities of belonging to a given auditory profile (AD). The relations between the AD_I_ and AD_II_ with the variables considered in the study were investigated using stepwise linear regression models. The variables included in the model fitting were the outcome variables resulting from the supra-threshold tests, except for AUD_LF_ and AUD_HF_, and listeners with a high probability of not being identified as any of the four profiles (P(U) > 0.5) were discarded. The criterion for adding a variable as a predictor of one of the distortions was an improvement of the adjusted *R*_2_ by more than 0.01.

### Decision trees

A decision tree was fitted to the entire dataset following the splitting criterion of weighted impurity (Breiman et al. 2017). Since it was of interest to obtain a decision tree with outcome measures beyond audiometry, the variables from the pure-tone audiometry were excluded from this analysis (i.e., AUD_LF_ and AUD_HF_). The resulting decision tree was pruned to only have three levels and a maximum of seven binary splits. Because of the missing data, the decision tree was surrogated, i.e., it ignored the missing data to facilitate its interpretability.

## Results

### Summary statistics of the Dataset

The percentiles of the outcome variables corresponding to the 75 participants and excluding the “optimal performer” are show in Table 2.

**Table 2.** Results of the test battery (BEAR3 dataset) presented as the 5th, 25th, 50th and 75th and 95th percentiles

| Variables | 5th | 25th | 50th | 75th | 95th | Unit |
| --- | --- | --- | --- | --- | --- | --- |
| $AUD_{LF}$ | 6.08 | 18.33 | 25.00 | 35.00 | 45.33 | dB HL |
| $AUD_{HF}$ | 22.41 | 45.83 | 54.16 | 62.08 | 70.58 | dB HL |
| $HTL_{LF}$ | 2.45 | 16.25 | 24.16 | 36.67 | 48.96 | dB HL |
| $HTL_{HF}$ | 16.87 | 35.62 | 48.75 | 57.91 | 71.29 | dB HL |
| FLFT | 4.12 | 6.27 | 7.94 | 10.24 | 12.20 | kHz |
| maxDS | 86.58 | 94.28 | 97.00 | 98.77 | 100.00 | % Corr. |
| $SRT_Q$ | 15.57 | 31.74 | 42.06 | 50.67 | 60.14 | dB HL |
| $SRT_N$ | -0.87 | 0.71 | 1.76 | 3.86 | 8.73 | dB SNR |
| $SS_{4dB}$ | 25.00 | 52.50 | 72.50 | 84.38 | 92.50 | % Corr. |
| $IPD_{FMAX}$ | 245.23 | 528.69 | 695.54 | 928.37 | 1109.20 | Hz |
| $BP_{20}$ | 15.00 | 71.25 | 95.00 | 100.00 | 100.00 | % Corr. |
| BMR | 7.50 | 12.79 | 15.75 | 17.62 | 22.25 | dB |
| $MCL_{LF}$ | 65.54 | 75.62 | 79.79 | 84.37 | 91.71 | dB HL |
| $MCL_{HF}$ | 66.37 | 74.37 | 80.21 | 88.95 | 98.21 | dB HL |
| $Slope_{LF}$ | 0.30 | 0.35 | 0.41 | 0.55 | 0.67 | CU / dB HL |
| $Slope_{HF}$ | 0.38 | 0.56 | 0.75 | 0.91 | 1.52 | CU / dB HL |
| $Dyn_{RLF}$ | 55.12 | 64.37 | 77.50 | 86.87 | 98.29 | dB |
| $Dyn_{RHF}$ | 34.12 | 41.25 | 50.20 | 61.66 | 82.79 | dB |
| $TiN_{LF}$ | 67.57 | 69.93 | 71.56 | 73.56 | 76.42 | dB HL |
| $TMR_{LF}$ | 3.32 | 6.81 | 7.92 | 10.12 | 12.00 | dB |
| $SMR_{LF}$ | 11.87 | 17.31 | 21.06 | 23.19 | 25.37 | dB |
| $TiN_{HF}$ | 70.03 | 72.12 | 73.93 | 75.87 | 79.26 | dB HL |
| $TMR_{HF}$ | 0.67 | 5.85 | 8.83 | 10.78 | 15.01 | dB |
| $SMR_{HF}$ | 4.21 | 12.68 | 21.62 | 27.21 | 33.56 | dB |
| $sSTM_8$ | -0.40 | 0.49 | 2.36 | 3.07 | 3.07 | d' |
| $fSTM_8$ | -10.71 | -7.90 | -5.68 | -1.50 | 0 | dB 20log(m) |

### Data-driven auditory profiling

The BEAR3 dataset was analyzed with an iterative data-driven auditory profiling method. The main results can be summarized by the probabilities of the listeners of belonging to a given auditory profile (A-D) and the expected performance of the listeners identified in each of the four groups (prototypes).

Figure 3 shows the results of the analysis where each listener is located in the two-dimensional space according to their degree of type-I and type-II distortion. The degree of distortion was calculated based on the probability of belonging to any of the four auditory profiles. Listeners located close to a corner exhibited a high probability of belonging to a corresponding profile. Uncategorizable listeners were placed inside the grey area representing a low probability of being classified as belonging to one of the four auditory profiles. After 1000 iterations, the probability of being uncategorizable P(U) was also calculated. If this probability was greater than the probability of belonging to any of the four profiles, the listener was considered “uncategorizable”. Profile A (n = 24) and Profile C (n = 22) represented the most populated groups. The five normal-hearing listeners were placed at the bottom-left corner in Profile A. Profiles B (n = 13) and Profile D (n = 9) represented smaller subgroups. Four listeners showed a high probability of being uncategorizable (labeled between asterisks in Figure 3), and four other listeners were “inconclusive” as reflected in similar probabilites of belonging to two profiles. The average results of the five listeners showing the highest probabilities of belonging to each one of the auditory profiles (excluding the normal-hearing listeners) were considered to represent the prototypes shown in Figure 4.

**Figure 3.**
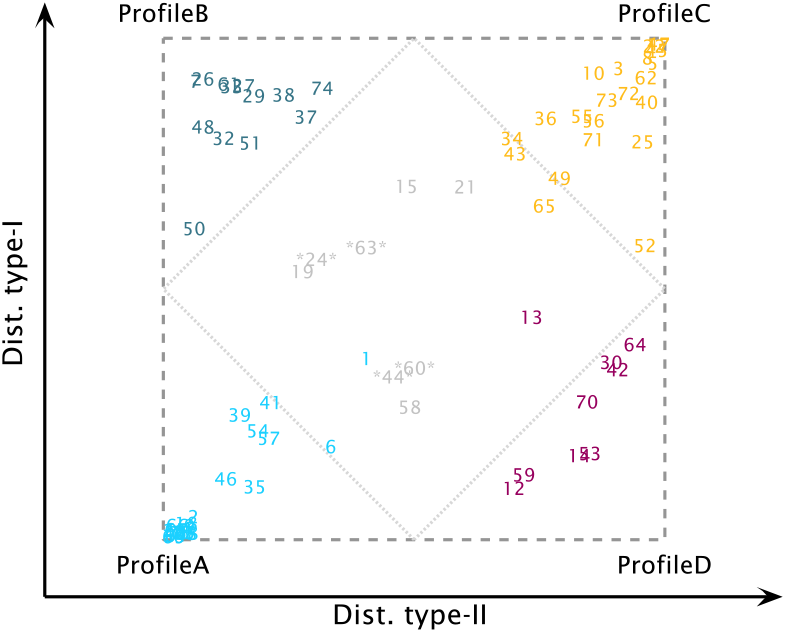
Square representation of the auditory profiles. The listeners are placed in the square representation based on their probability of belonging to one of the subgroups. The inner rhombus delimits the area of inconclusive profile membership, i.e., listeners showing a probability < 0.5 of belonging to any subgroup. The listeners marked with two asterisks were considered uncategorizable and showed P(U) > 0.5..

**Figure 4.**
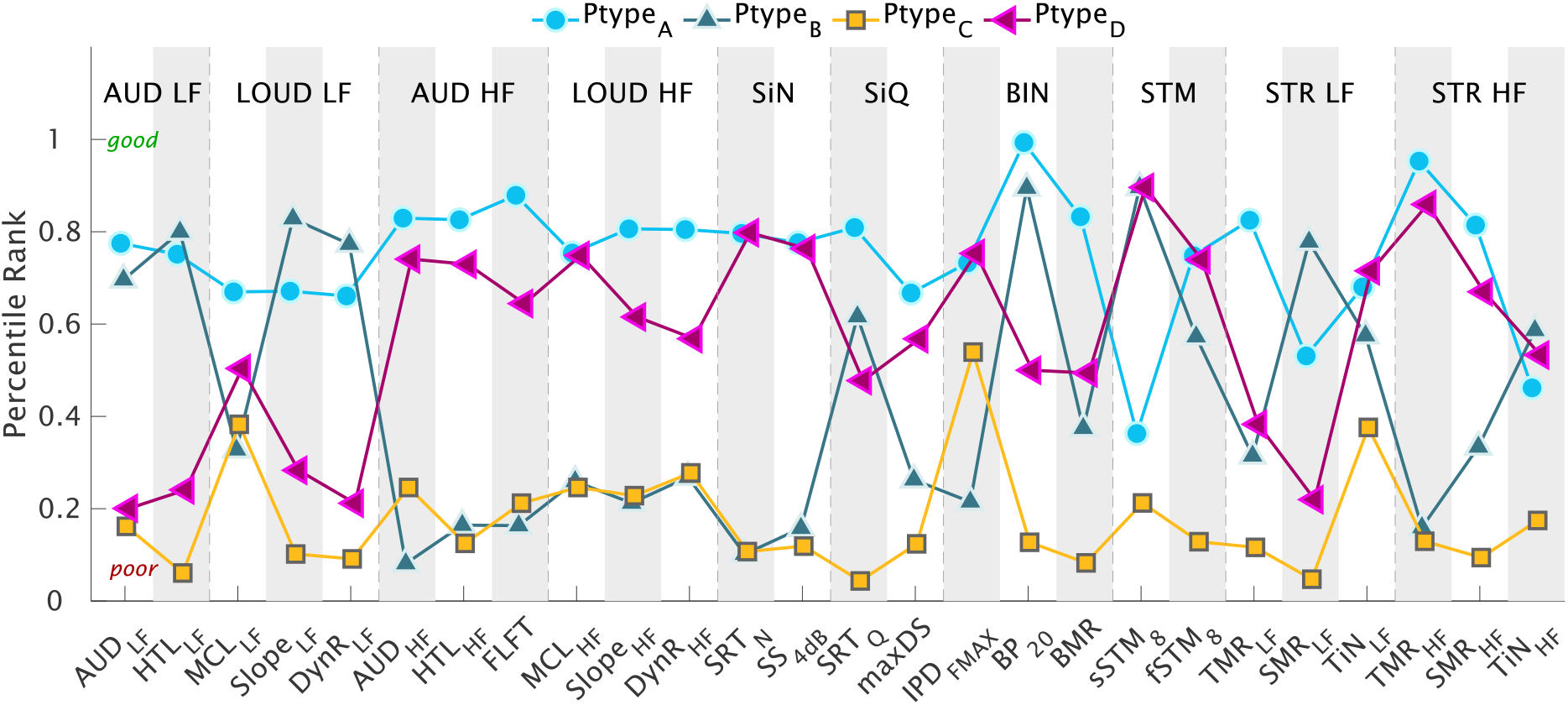
Prototypes (Ptype): Percentile rank across variables corresponding to the extreme exemplars of the different patterns found in the data. The 26 outcomes corresponding to the different aspects of auditory processing are divided into the following subdimensions: AUD: Audibility, LOUD: Loudness, SiN: Speech-in-noise perception, SiQ: Speech-in-quiet, BIN: Binaural processing abilities. STM: Spectro-temporal modulation sensitivity, STP: Spectro-temporal processing abilities, divided into temporal and spectral masking release as well as tone in noise detection. Subgroups of measures with frequency-specific outcomes were divided into low (LF) and high (HF) frequencies.

The prototypes show archetypal patterns in the data associated with the performance obtained by the four different groups. A higher percentile rank corresponds to a higher percentile of the overall data distribution and thus to a “good” performance. Each point in Figure 4 corresponds to the mean of the listeners forming the corresponding prototype. Likewise, a low percentile rank corresponds to a “poorer” performance. Prototype A (blue circles in Figure 4) showed a good performance in most of the outcome measures. However, the outcomes sSTM_8_ and TiN_LF_ were below the 50th percentile. Prototype C (yellow squares) showed the poorest performance for most outcome measures, with only MCL_LF_ and IPD_fmax_ above the 30th percentile. Prototype B (dark-green upwards-pointing triangles), with a high degree of distortion type-I and a low degree of distortion type-II, showed a good performance for the outcome measures obtained at lower frequencies and for BP_20_, whereas performance was poor for the outcomes obtained at higher frequencies, IPD_fmax_ and for the speech-in-noise perception tests. In contrast, prototype D (magenta left-pointing triangles), with a high-degree of distortion type-II and a low degree of distortion type-I, showed a poor performance for outcome measures obtained at low frequencies, especially in terms of loudness, TMR_LF_ and SMR_LF_, whereas the performance was good (above the 60th percentile) for most outcomes measures obtained at higher frequencies, speech-in-noise perception and IPD_fmax_. The prototypes showed opposite results for the profiles located in opposite corners of Figure 3 (A vs. C and B vs. D).

### Relations between auditory distortion types and outcome measures

The relations between the two types of distortions and outcome measures were studied using stepwise regression analysis (Table 3). Distortion type-I was found to be associated with elevated hearing thresholds at higher frequencies, a reduced temporal masking release and increased tone-in-noise detection thresholds at low frequencies. Furthermore, distortion type-I was significantly correlated with SRT_N_ (*r* =0.76; *p<* 0.0001), even when the effects of audibility were partialled out (*r* =0.33; *p<* 0.01). In contrast, the correlations found between distortion type-I and speech recognition in quiet (*r* =0.71; *p<* 0.0001) were not significant when partialling out audibility (*r* =0.15; *p>* 0.1). Distortion type-II was only associated with hearing thresholds at low frequencies. The restrictive criterion (increase of *R*_2_ *>* 0.01) did not include other variables in the model. However, distortion type-II was significantly correlated with the slope of the loudness function (*r* =0.72; *p<* 0.0001) and with the amount of spectral masking release at low frequencies (*r* =0.61; *p>* 0.0001). In addition, distortion type-II was correlated with SRT_Q_ (*r* =0.83; *p<* 0.0001) but not with SRT_N_ (*r* =0.21; *p>* 0.05). However, the correlation between SRT_Q_ and distortion type-II was weaker when controlling for the effects of audibility (*r* =0.30; *p<* 0.05). Moreover, the majority of the auditory outcomes were not significantly correlated with distortion type-II when hearing thresholds were partialled out, except for TMR_HF_(*r* =0.35; *p<* 0.01).

**Table 3.**
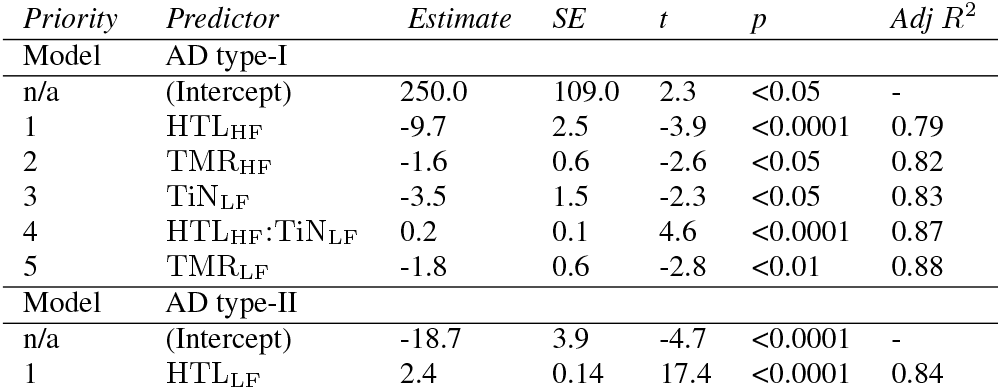
Stepwise regression analysis of auditory distortion (AD) type-I and type-II. The priority was established based on the accumulated adjusted *R*^2^ > 0.01. Columns show the predictor name, the estimate, standard deviation (SE), t-value and probability of a significant contribution (p).

The outcome measures related to binaural processing abilities (Figure 4) gave unexpected results. Indeed, the prototypes showed opposite trends for IPD_fmax_ and BP_20_, which could indicate that they reflect different auditory distortions. Distortion type-I was significantly correlated with both IPD_fmax_ and BP_20_, but only BP_20_ remained significant after controlling for audibility (*r* = −0.36; *p<* 0.01). In contrast, distortion type-II was only correlated with BP_20_ (*r* = −0.58; *p<* 0.0001) before partialling out the effects of audibility (*r* = −0.1; *p* =0.5). Besides, IPD_fmax_ was neither correlated with any of the two distortion types when controlling for audibility nor with any of the other binaural processing abilities outcome measures (*r <<* 0.1; *p>* 0.15). Instead, IPD_fmax_ was highly correlated with the tone-in-noise detection threshold at low frequencies (*r* = −0.53; *p<* 0.0001) — one of the main predictors of distortion type-I — even when audibility was partialled out (*r* = −0.56; *p<* 0.0001).

### Decision tree for the identified auditory profiles

Figure 5 shows the decision tree fitted to the BEAR3 dataset using the identified auditory profiles as well as the uncategorizable listeners. The decision tree has three levels. The first level corresponds to high-frequency hearing loss as estimated using ACALOS, which splits the listeners into two branches: Profiles A and D (HTL_HF_ < 49 dB HL) are separated from Profiles B and C (HTL_HF_ > 49 dB HL), together with one listener from Profile D. Thus, this first level makes a classification based on the degree of distortion type-I. The second level corresponds to outcomes measured at low frequencies and estimated using the loudness functions, which divide the listeners according to their degree of distortion type-II. Profile D (HTL_LF_ > 28 dB HL) and Profile C (Slope_LF_ > 0.4 CU/dB and maxDS < 100%). The third level makes use of outcomes related to loudness, spectro-temporal modulation and spectral masking release for classifing the uncategorizable listeners.

**Figure 5.**
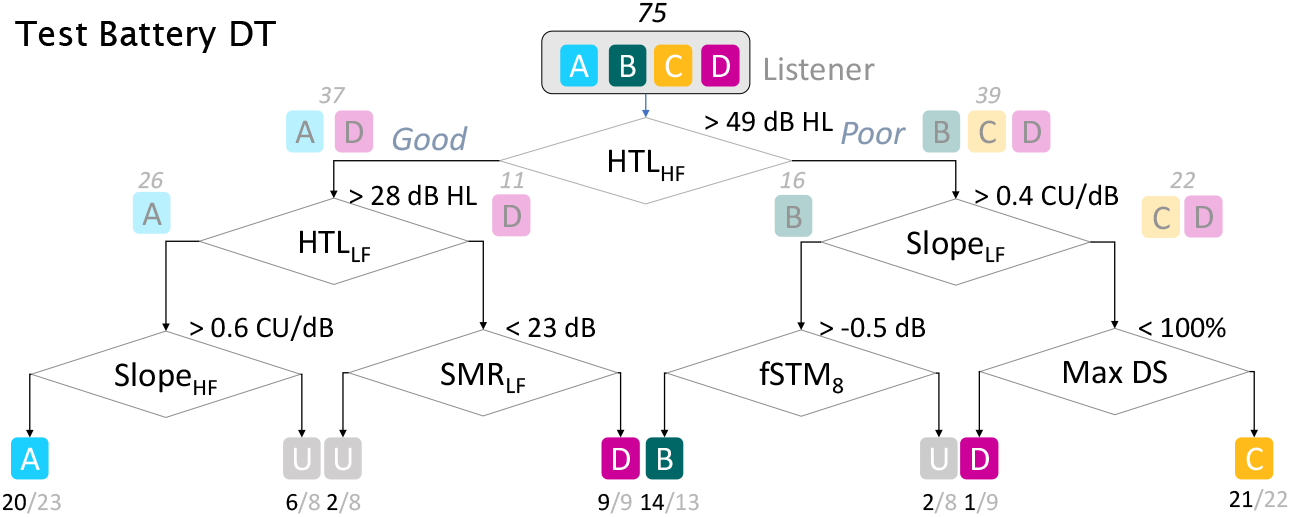
Decision tree fitted to the dataset using the auditory profiles as the output. For each binary split, the right branch corresponds to a “poor” result and the left branch to a “good” result. In each binary split, the number of listeners assigned to each branch are shown together with the most likely outputs. The classes (A-D) are together with the number of listeners belonging to that class and the number of identified listeners for a given profile.

## Discussion

The data-driven method for auditory profiling presented here provides new knowledge about hearing loss characterization. Regarding previous data-driven auditory profiling (Sanchez-Lopez et al. 2018), the present results are in good agreement with the analysis performed on the data of Johannesen et al. (2016) data set. This suggests that the use of data from a representative sample of different degrees of hearing loss (e.g. in Johannesen et al. 2016) and a normal-hearing reference (e.g. in Thorup et al. 2016) is crucial for robust profile-based hearing-loss characterization.

### Two types of distortion to characterize individual hearing loss

The term “distortion” in hearing science has typically been associated with elevated SRT_N_, as reflected in Plomp’s SRT model (Plomp 1978). Here, we introduced the term “auditory distortions” to describe the perceptual consequences of sensory hearing impairment, including (but not limited to) loss of sensitivity. The two types of perceptual distortions considered here should thus be considered as consequences, and not sources of, sensory impairments. An interesting aspect of our data-driven profiling method is that the auditory distortions reflect two fairly independent dimensions of perceptual deficits associated with sensorineural hearing impairments. To reiterate, distortion type-II was associated with low-frequency hearing loss and steep loudness functions. However, listeners with a high degree of distortion type-II and a low degree of distortion type-I (Profile D) did not exhibit exclusive audibility loss, as they also exhibited an abnormal loudness growth and a reduced spectral masking release. Distortion type-I was associated with elevated hearing thresholds at higher frequencies and was significantly correlated with elevated SRT_N_. Furthermore, for this distortion type, TMR_HF_ and TiN_LF_ were poorer even when the effect of the audiometric thresholds was controlled for.

Although Plomp’s attenuation and distortion components are often assumed to be independent, some impairment mechanisms may, in fact, affect both speech-in-noise perception and audiometric thresholds, especially at high frequencies (Moore 2016), which is consistent with distortion type I. Schädler et al. (2020) attempted to model supra-threshold auditory deficits that are independent of audibility loss. Their results suggested that reduced speech intelligibility represents an auditory perceptual deficit that may be associated with reduced tone-in-noise detection which is in agreement with the results from the current study. However, as demonstrated here, speech-in-noise perception can also be affected by deficits that covary with audiometric thresholds (distortion type-I), which should not be underestimated, especially when the high-frequency hearing loss exceeds 50 dB HL (Profiles B and C), as depicted in Figure 6.

**Figure 6.**
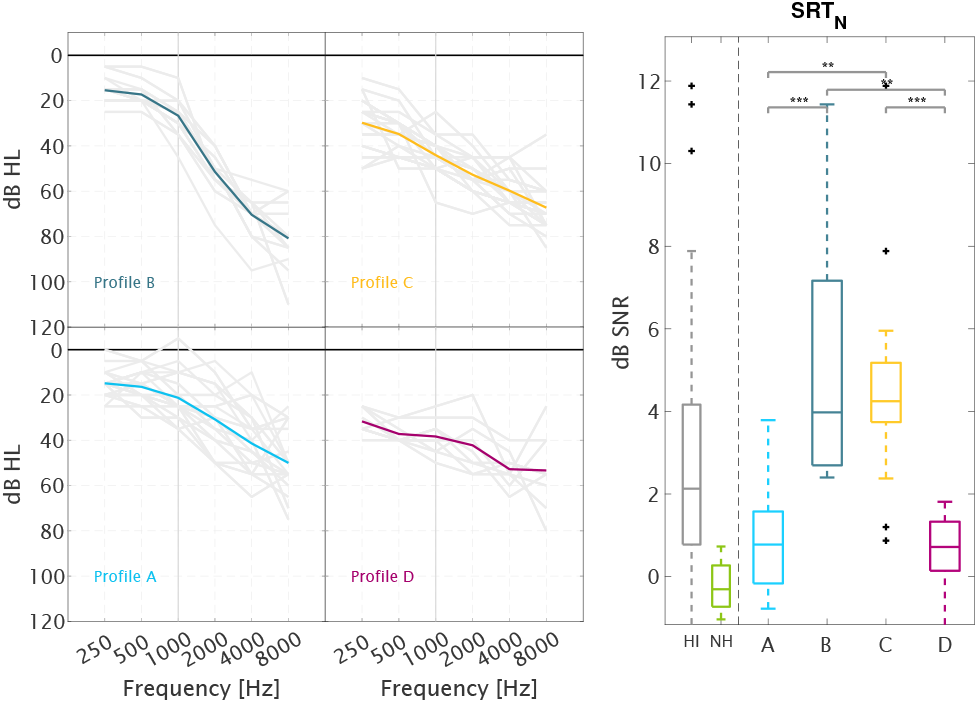
Audiometric thresholds of the four auditory profiles and speech intelligibility in noise. Left panel: The average audiometric thresholds of each profile are shown together with the audiograms of the individual ears. Right panel: Speech reception thresholds in noise (SRTN), with boxplots of the HI and NH data (left) and the four auditory profiles (right). The multicomparison analysis revealed significant differences between the groups (*** p < 0.0001, ** p < 0.001).

Regarding the ‘neural component’ associated with reduced binaural processing abilities (Kollmeier and Kiessling 2018), the BIN measures considered in the present study provided contradictiory results in connection to the proposed auditory profiles. Even though IPD_fmax_ represents a test that has been proposed to reveal binaural disabilities related to the disruption of temporal fine structure (TFS) coding (Füllgrabe and Moore 2017). Recent studies have linked the detection of interaural phase differences to outcomes from cognitive tests (Strelcyk et al. 2019; Füllgrabe et al. 2015). This suggests that IPD_fmax_ might not reflect a purely auditory process but might also depend on top-down processes such as processing speed or selective attention. Since IPD_fmax_ and TiN_LF_ were strongly correlated, the two tasks might be affected by either cognitive or auditory processes, which should be investigated further.

The two types of auditory distortions shown here were consistent with Plomp’s approach (Plomp 1978). The profiles with a low degree of distortion type-I (Profiles A and D) exhibited a loss of sensitivity but their speech-reception thresholds were comparable to the ones of NH listeners. In contrast, the profiles with a high degree of distortion type-I (Profiles B and C) exhibited elevated SRTs (see the right panel of Figure 6). However, the two auditory distortion types presented here are, in fact, the result of a data-driven analysis of a large multi-dimensional dataset rather than the conceptual interpretation of speech intelligibility deficits. Distortion type-I may then be considered as a “speech intelligibility related distortion” and distortion type-II as a “loudness perception related distortion”. Nevertheless, the listeners with higher degrees of the two types of distortions showed perceptual deficits with respect to spectro-temporal processing and binaural processing abilities thus reflecting deficits that are beyond a simple combination of loudness and speech-intelligibility deficits.

### Auditory profiles and hearing-loss phenotypes

Figure 6 shows the average audiometric thresholds corresponding to the listeners belonging to the four robust auditory profiles. Profile A corresponds to a mild, gently sloping high-frequency hearing loss; Profile B corresponds to a steeply sloping high-frequency hearing loss; Profile C corresponds to a low-frequency hearing loss between 30 and 50 dB HL and above 50 dB HL at high frequencies; and Profile D corresponds to a fairly flat hearing loss with audiometric thresholds between 30 and 50 dB HL. Interestingly, these four audiometric configurations look similar to the audiometric phenotypes (Dubno et al. 2013), which are based on Schuknecht’s metabolic and sensory types of presbyacusis (Schuknecht and Gacek 1993). The main difference between the two approaches is that the audiometric threshold functions shown here correspond to four subgroups of HI listeners, which are the result of a data-driven analysis involving various auditory measures (and not only audiometric thresholds). Based on audiometric thresholds only, the listeners in Profile A and Profile B would be classified into the same phenotypical category (i.e. sensory hearing loss according to Dubno et al.), even if they present substantial differences in supra-threshold auditory hearing abilities such as speech intelligibility.

In previous studies, metabolic hearing loss (MHL) yielded flat elevated audiometric thresholds, but did not affect speech intelligibility in noise (Pauler et al. 1986), which is consistent with the results of the present study for Profile D listeners. In MHL, the atrophy of the stria vascularis produces a reduction of the EP in the scala media (Schmiedt et al. 2002). The EP loss mainly affects the electromotility properties of the OHC (i.e. the cochlear amplifier). Therefore, metabolic hearing loss can be considered as a cochlear gain loss that impairs OHC function across the entire cochlea. This, in turn, affects the hearing thresholds and is associated with a reduced frequency selectivity (Henry et al. 2019). In the present study, Profile D was characterized by an abnormal loudness function, particularly at low frequencies, and a significantly reduced spectral masking release, although speech-in-noise intelligibility and binaural TFS sensitivity were near-normal. However, one needs to bear in mind that the results observed for the listeners in Profile D might also be compatible with other types of impairments. Sensory hearing loss (SHL) is typically associated with OHC dysfunction, which yields elevated thresholds at more specific frequency regions, a loss of cochlear compression and reduced frequency selectivity (Ahroon et al. 1993). However, audiometric thresholds above about 50 dB HL at high frequencies cannot be attributed only to OHC due to the limited amount of gain induced by the OHC motion, which implies additional IHC loss or a loss of nerve fibers (Wolak et al. 2019; Hamernik et al. 1989; Stebbins et al. 1979). Therefore, listeners classified as Profile B or Profile C (i.e. with a higher degree of distortion type-I and a high-frequency hearing loss) may exhibit a certain amount of IHC dysfunction that might produce substantial supra-threshold deficits. Animal studies have shown that audiometric thresholds seem to be insensitive to IHC losses of up to about 80% (Lobarinas et al. 2013). This suggests that hearing thresholds > 50 dB HL might indicate the presence of hearing deficits that may distort the internal representation, not only in terms of frequency tuning but also in terms of a disruption of temporal coding due to the lack of sensory cells (Stebbins et al. 1979; Moore 2001).

Profile B’s audiometric thresholds are characterized by a sloping hearing loss with normal values below 1 kHz. However, Profile B exhibited the poorest performance in the IPD_fmax_ test, which cannot be explained by an audibility loss. Neural presbyacusis is characterized by a loss of nerve fibers in the spiral ganglion that is not reflected in the audiogram. Furthermore, primary neural neurodegeneration, recently termed cochlear synaptopathy (Kujawa and Liberman 2009; Wu et al. 2019) or deafferentation (Lopez-Poveda 2014), might be reflected in the results of some of the supra-threshold auditory tasks used here. However, the perceptual consequences of primary neural degeneration are still unclear due to the difficulty of assessing auditory nerve fibers loss in living humans (Bramhall et al. 2019). This makes it difficult to link the effects of deafferentation to the reduced binaural processing abilities observed in listeners in Profile B and Profile C.

As suggested in Dubno et al. (2013), the audiometric phenotype characterized by a severe hearing loss (similar to the one corresponding to Profile C) might be ascribed to a combination of MHL and SHL. In the present study, Profile C listeners performed similarly to Profile B listeners in supra-thresholds tasks related to distortion type-I (e.g. SRT_N_ and TMR_HF_) and also similarly to Profile D listeners in tasks related to distortion type-II (e.g. loudness perception). In contrast, Profile C listeners also showed poorer performance in tests such as binaural pitch detection, tone-in-noise detection and spectro-temporal modulation sensitivity, which is not consistent with the idea of a simple superposition of the other profiles. As mentioned above, these deficits observed in Profile C listeners might be a consequence of auditory impairments that are unrelated to the loss of sensitivity, such as deafferentation, which can be aggravated by the presence of MHL and SHL. However, it has been found that spectrotemporal modulation sensitivity could be a good predictor of aided speech perception only in the cases of a moderate high-frequency hearing loss (Bernstein et al. 2016). They suggested that cognitive factors might be involved in the decreased speech intelligibility performance when the high-frequency hearing loss is >50 dB HL. Therefore, Profile C listeners might be affected by both auditory and non-auditory factors that worsen their performance in some demanding tasks.

### Stratification in hearing research and hearing rehabilitation

In the present study, the two principal components of the dataset seemed to be dominated by the listeners’ low-and high-frequency hearing thresholds. This suggests that supra-threshold deficits might be associated with different forms of audibility loss. However, other supra-threshold hearing deficits, which do not covary with a loss of sensitivity, might be hidden in the four auditory profiles and could explain the individual differences across listeners belonging to the same profile. To explore these “additional deficits” not covered by the present approach, stratification of the listeners might be necessary. Lőcsei et al. (2016) investigated the influence of TFS on speech perception for different interferers. In their study, the HI listeners were divided into groups based on the degree of hearing loss at high frequencies. In their study, stratification of the listeners into two subgroups helped reduce the potential effect of audibility on speech intelligibility. In another study, Papakonstantinou et al. (2011) studied the correlation of different perceptual and physiological measures with speech intelligibility in stationary noise. In their study, all the listeners had a steeply sloping high-frequency hearing loss consistent with Profile B. Both studies (Lőcsei et al. 2016; Papakonstantinou et al. 2011) included measures of frequency discrimination thresholds and speech-in-noise perception. However, Papakonstantinou et al. (2011) tested a larger group of HI listeners with fairly similar audiograms in only one speech condition that led to a highly significant correlation between frequency discrimination and speech intelligibility in stationary noise. This suggests that the stratification of the listeners and the investigation of certain phenomena in separated auditory profiles might reveal new knowledge about hearing impairments that are not generalized to the entire population of HI listeners.

Other approaches have attempted to identify why listeners with similar audiograms present substantial differences in suprathreshold performance. Recently, Souza et al. (2020) showed how older HI listeners vary in terms of their ability to use specific cues (either spectral or temporal cues) for speech identification. Their results showed a so-called “profile cue” that characterizes the listener’s abilities in terms of spectro-temporal processing. Some listeners utilized temporal envelope cues and showed good temporal discrimination abilities, whereas other listeners relied on spectral cues and were able to discriminate spectral modifications in a speech signal. The “profile cue” resulted from a syllable identification task, which was independent of the audiometric thresholds and associated with the spectral discrimination task. Even though the spectral discrimination task was promising for predicting the “profile cue", this test seemed to be influenced by the audiometric thresholds. Since their participants presented audiograms similar to the ones observed for Profiles B, C and D (Figure 6), it is possible that the categorization of the listeners based on auditory profiling, together with the spectral discrimination task (not included in the present study), could enable an efficient prediction of the “profile cues” and clarify its connection to supra-threshold auditory deficits.

Sensory rehabilitation of the hearing deficits involves the use of hearing devices. The criteria for candidacy of implantable technology (Kirkby-Strachan and Que-Hee 2016; Irving et al. 2014), which are based on the benefit observed by using acoustical devices, are sometimes insufficient. New findings with electro-acoustic stimulation (Imsiecke et al. 2019) suggest that this technology might benefit patients with a high degree of speech-intelligibility related deficits, i.e. with HL_HF_ *>* 50 dB HL as shown here. Despite the detrimental effect of the interaction between electrical and acoustical stimulations, Imsiecke et al. (2019) showed that the use of masking-adjusted fittings with the aim of reducing electric-on acoustic masking strength was beneficial for speech intelligibility even in the cases with good residual hearing. Therefore, the present characterization into auditory profiles might support a revision of the candidacy of implantable devices that might include less severe hearing losses at high frequencies. Furthermore, auditory profiling showed potential for hearing diagnostic that can help disentangle the effects of different types of impairments. This might be particularly useful for the development of therapeutics for hearing loss (Kujawa and Liberman 2019) supporting precision medicine.

Overall, the participants of the present study would be candidates for hearing aids. Hearing-aid users often show a large variability in terms of benefit and preference to specific forms of hearing-aid processing (Neher et al. 2016; Picou et al. 2015; Souza et al. 2019). In some studies, the HI listeners were stratified based on their audiograms (Gatehouse et al. 2006; Keidser et al. 1995; Keidser and Grant 2001; Larson et al. 2002). However, the existing hearing-aid fitting rules do not make use of supra-threshold auditory measures that might help tune the large parameter space of modern hearing technology. In fact, the HA parameters are still adjusted based on the audiogram and empirical findings that provide some fine-tuning according to the HA user experience or the gender of the patient (Keidser et al. 2012). Furthermore, candidacy for specific hearing-aid processing (e.g. beamforming) could be driven by specific supra-threshold auditory deficits (e.g., Neher et al. 2017; Füllgrabe et al. 2018), or non-auditory aspects (e.g., Neher et al. 2016; Souza et al. 2019). The four auditory profiles presented here showed significant differences in supra-threshold measures related to two independent dimensions, a “speech intelligibility-related distortion” and a “loudness perception-related distortion”. Therefore, profiling method allows stratifying the listeners beyond what can be achieved with an audiogram. This may help optimize hearing-aid fitting parameters for a given patient. Recently, it has been suggested that different advanced signal processing strategies should be considered to compensate for different cochlear pathologies (Henry et al. 2019). Since the four auditory profiles showed interesting similarities to the sensory and metabolic phenotypes proposed by Dubno et al. (2013) new forms of signal processing aimed at overcoming the hearing deficits associated with the two identified dimensions, may be developed and evaluated. Furthermore, the current approach may inspire different forms of model-based hearing loss compensation (Bondy et al. 2004) to restore auditory function based on biologically inspired technology. This can lay the foundations for precision medicine (Jameson and Longo 2015) applied to the perceptual rehabilitation of the hearing deficits. The implementation of a clinically feasible classification procedure depends on different factors, such as time efficiency and the availability of the necessary auditory tests and hearing-aid algorithms.

### Limitations of the data-driven auditory profiling approach

A clear limitation of the data-driven method proposed here is its constraint to two dimensions of independent auditory deficits and four subgroups. The advantage of the proposed method is that it can provide results that can be easily interpreted even when using advanced computational methods for data analysis. Usually, advanced data-driven methods provide meaningful results, but they are not necessarily linked to an initial hypothesis. In our approach, the imposed link between method and hypothesis leads to a constraint in terms of dimensions and subgroups. However, it would be interesting to extend the current data-driven method to allow for a third (or even higher) dimension which might partly explain additional variance in the data in future research.

The definition of the auditory profiles reflected the main sources of hearing deficits in a relatively large and heterogeneous population of HI listeners. However, this group only contained older adults (>60 years) with symmetric sensorineural hearing losses. An extension of the auditory profiling method proposed here might be based on an even more heterogeneous group of participants, which might require different data-analysis techniques for proper analysis and interpretation (Hinrich et al. 2016). The insights from the current method could then be applied mainly to a population of mild-to-severe age-related hearing losses and to some extent to other types of non-syndromic hearing losses, e.g. noise-induced hearing loss, but cannot be generalized to the whole variability of existing auditory pathologies.

The presented data-driven approach for hearing loss characterization was intentionally limited to the use of psychoacoustical measures and the use of auditory tests with potential for clinical implementation. Physiological measures, such as otoacoustic emissions and auditory evoked potentials, were not considered in the current approach. This was a decision in the interest of the characterization of the perceptual consequences derived of the hearing deficits rather than the “sources” of the hearing loss. Cognitive factors are also important for characterizing the overall “listening profile” of individuals with hearing loss, as suggested in several studies (e.g., Rönnberg et al. 2016; Humes 2007). In the present study, cognitive factors were considered as a confound rather than a missing part of the auditory profiling approach. A better understanding of the sensory dysfunction is needed to provide an efficient compensation of the hearing deficits rather than a compensation for the audibility loss. However, it would be of great interest to explore the cognitive factors from a bidirectional point of view. Cognition can affect the perception of the auditory stimuli presented in the test battery and the listener’s cognitive resources can also be affected by the “distortions” reflected by the auditory profiles and lead to an effortful listening experience (Pichora-Fuller et al. 2016; Peelle 2018).

Besides the potential for clinical implementation, the tests that were language independent were prioritized. However, a test battery representing speech intelligibility deficits would be of great relevance. Such a test battery could be tested on a population of people with different hearing abilities and analyzed using a similar data-driven profiling method as the one presented here. This test battery might involve speech intelligibility tests in the presence of different interferers and spatial configurations (e.g. Lőcsei et al. 2016). Besides, it might contain tests where speech intelligibility is affected by reverberation, audible distortions, or the use of amplification. In such a study, phenomena such as masking release or binaural unmasking could be further investigated using a data-driven approach.

The current study focused on the basis of auditory profiling and not on their clinical application. For the latter, other considerations such as the reliability, time-efficiency and ease of administration of the tests must be taken into account. In its present form, the BEAR test battery may be unfeasible to be administered in the clinic, but some individual tests have the potential to be implemented in the clinical practice and to guide the characterization of individual hearing deficits. The evaluation and optimization of the auditory profiling approach should be undertaken carefully, bearing in mind that the purpose of the classification is a better intervention. First, it needs to be demonstrated that the stratification applied to hearing rehabilitation is beneficial for the patient, and second, the identification of a set of measures to better characterize the auditory deficits needs to be optimized to find a balance between the time spent in the additional tests and the benefit obtained with this approach.

### Conclusion

Using a data-driven approach, four auditory profiles (AB-C-D) were identified that showed distinct differences in terms of supra-threshold auditory processing capabilities.. The listeners’ hearing deficits could be characterized by two independent types of auditory distortion, a “speech intelligibility-related distortion” affecting listeners with audiometric thresholds >50 dB HL at high frequencies, and a “loudness perception-related distortion” affecting listeners with audiometric thresholds >30 dB HL at low frequencies. The four profiles showed similarities to the audiometric phenotypes proposed by Dubno et al. (2013), suggesting that Profile B may be resulting from a sensory loss and Profile D may be resulting from a metabolic loss. Profile C may reflect a combination of a sensory and metabolic loss, or a different type of hearing loss that results in substantially poorer supra-threshold auditory processing performance. The success of this approach provides new methods to identify homogeneous sub-populations to better investigate the perceptual consequences of different etiologies. The current results enable “precision audiology” and provide new avenues for developing auditory-profile based compensation strategies for hearing rehabilitation.

## Data Availability

The data used in the study is publically available in a Zenodo Repository:
Sanchez-Lopez, R., Nielsen, S. G., El-Haj-Ali, M., Bianchi, F., Fereckzowski, M., Canete, O., … Santurette, S. (2019). Data from ''Auditory tests for characterizing hearing deficits: The BEAR test battery.'' https://doi.org/10.5281/zenodo.3459579

https://zenodo.org/record/3459580

## Supplemental material

Additional figures illustrating the performance of the auditory profiles in each individual tests are provided as supplemental files. The figures are presented in boxplots as the right panel of Figure 6. The correlations and partial correlations of each of the variables with the estimate of auditory distortions type-I and type-II are also provided as additional files.

## Acknowledgements

We thank F Bianchi, SG Nielsen, M El-haj-Ali, OM Cañete, and M Wu for their contribution to the design of the BEAR test battery, and work in the data collection. Also, we appreciate the valuable input from JB Nielsen, J Harte, W. Whitmer, G Naylor and O. Strelcyk among others. We would also like to thank G Encina-Llamas, AA Kressner and EN MacDonald who suggested some improvements in an earlier version of this manuscript. This work was part of the Better hEAring Rehabilitation project, the funding and collaboration of all partners are sincerely acknowledged.

## Footnotes

* Thorup et al. (2016)’s dataset was collected in a clinical setup using listeners with either near-normal audiometric thresholds (26 listeners), obscure dysfunction (4 listeners) or mild-to-moderate high-frequency hearing loss (29 listeners). The age of the listeners ranged from 41 to 70 years in the near-normal hearing group and from 52 to 80 years in the hearing-impaired group. The dataset contained 27 variables consisting of: 1) audiometric thresholds, 2) loudness perception, 3) speech perception in quiet and in noise, 4) binaural processing abilities and 5) the reading span test.

† Johannesen et al. (2016)’s dataset was obtained in a research setting using 67 hearing-impaired listeners with moderate-to-severe hearing losses. The age of the listeners ranged from 25 to 82 years. The dataset contained 11 variables consisting of: 1) audiometric thresholds, 2) aided speech recognition thresholds, 3) frequency modulation detection and 4) basilar membrane compression estimates.

‡ Only one listener presented a pure-tone audiometric threshold above 70 dB HL at 2 kHz (listener 20).

